# Participation in a voluntary alcohol abstinence program (Alcohol Free for 40) with tracking of laboratory values and physical metrics improves biometric measures of health; a three-year retrospective analysis

**DOI:** 10.1101/2022.06.15.22276444

**Authors:** Amy K. Feehan, Megan B. Knapp, Jala Lockhart, Erin E. Arceneaux, Savanna Latimer, Anna Walter, Brittany N. Craft, Yvette P. Quantz, Hope Frugé, Maria Sylvester Terry, David Galarneau, Molly Kimball

**Affiliations:** Ochsner Clinic Foundation, 1514 Jefferson Highway, New Orleans, LA, USA 70121; The University of Queensland, Ochsner Clinical School, 1514 Jefferson Highway, New Orleans, LA, USA 70121; Xavier University of Louisiana, 1 Drexel Drive, New Orleans, LA 70125; The University of Alabama, Tuscaloosa, AL 35487

**Keywords:** community-based program, alcohol consumption, biometric tracking, temporary alcohol abstinence

## Abstract

**Background:** Laboratory and physical metric data were available from participants of Alcohol Free for 40 (AFF40), a 40-day alcohol abstinence program in Louisiana. This study was performed to determine whether biometric changes were detectable after 40 days.

**Materials and Methods:** This was a retrospective data review of three cohorts of AFF40 program participants. Participant pre- and post-program data were analyzed (n=113, 2019; n=105, 2020; n=344, 2021). The intervention was self-directed, 40-day alcohol abstinence with social support. Changes in liver enzymes, cholesterol, vitamin B12, weight, body fat, and blood pressure were measured. Paired t-tests were used to compare pre- and post-program metrics.

**Results:** Retention rates for participation were 63% (113/179; 2019), 23% (105/449; 2020), and 86% (344/400; 2021), despite the SARS-CoV-2 pandemic in 2020 and 2021. Maximal average changes were - 9 mmHg in systolic blood pressure, -4.3 mmHg in diastolic blood pressure, and -5.7 pounds of weight (all p<0.0001). Liver enzymes decreased; alanine aminotransferase decreased by 5.6 U/L, aspartate transaminase by 4.8 U/L, and gamma glutamyl transferase by 8.7 U/L (all p<0.0001). Vitamin B12 increased by 54.6 pg/mL (p<0.01). Total cholesterol dropped by 15.8 mg/dL (p<0.0001).

**Conclusion:** Participants in abstinence programming with social support improved liver enzymes, cholesterol, body fat, and other factors during a 40-day challenge.

**Highlights:**

- Programs for voluntary abstinence from alcohol have become popular around the world (Dry January, Sober October)
- Alcohol Free for 40, a post-Mardi Gras abstinence program, revealed significant reductions in blood pressure, weight, fat, liver enzymes, total cholesterol, and increased vitamin B12 in participants after just 40 days
- Program retention was high except in 2020 due to the SARS-CoV-2 pandemic
- Dieticians and other healthcare practitioners can replicate Alcohol Free for 40 for individuals or groups who want to improve metrics described in this study

## Introduction

In the United States, alcohol consumption is prevalent with more than half of adults consuming alcohol every month.^1^ Because of its molecular structure, alcohol is adept at entering cells and can have deleterious effects on almost all organ systems, with long-term consequences including high blood pressure, heart disease, stroke, liver disease, cancers, weakened immune system, memory problems, mental health issues, social consequences, and alcohol use disorders.^2-4^ Abstaining from alcohol, even for temporary periods, can have positive health effects. In two studies of alcohol consumers, abstinence periods of one month of alcohol abstinence resulted in improvements in liver enzyme levels, insulin sensitivity, weight, blood pressure, and cancer-related growth factors.^5, 6^

This study retrospectively reviewed physiological markers of health of participants who completed Alcohol Free for 40 (AFF40, www.alcoholfreefor40.com), an annual, community-based 40-day alcohol abstinence program. We hypothesize that participants who completed the program improved in several markers of health in a pooled analysis including liver enzymes, cholesterol, B12, weight, body fat, and blood pressure.

## Materials and Methods

### Design

Data was retrospectively collected from the electronic medical record of participants in the 2019, 2020, and 2021 cohorts of AFF40. Participants who completed both baseline (pre-challenge) and +40 days (post-challenge) laboratory and physical metrics were included in analysis. If a laboratory test was invalid or outside the limit of detection, pre- and post-values were excluded. This research was determined exempt by the Ochsner Clinic Foundation Institutional Review Board (FWA00002050).

### Sample

Over three years (2019-2021), enrollment was 179, 449, and 400, respectively and retention (completion of pre- and post-challenge labs) was 63% (113/179; 2019), 23% (105/449; 2020), and 86% (344/400; 2021). Physical metrics were only available in 2019 (n=83) and 2021 (n=304) due to the SARS-CoV-2 pandemic. In 2020, pre- and post-challenge labs were separated by 83 days rather than 40. Most participants were female (70-72%) and white (81-89%). The average age was 52.4 in 2019, 53.5 in 2020, and 53.7 in 2021.

### Measures

Analyzed laboratory data included a comprehensive metabolic panel (aspartate transaminase (AST), alanine aminotransferase (ALT), gamma glutamyl transferase (GGT)), lipid panel (including triglycerides), and vitamin B12 levels. Physical data was measured using the InBody570 (InBody Co., LTD, South Korea) and included weight, percent body fat, and pounds of body fat. Blood pressure was recorded from the iHealth Track Connected Blood Pressure Monitor (iHealth, Mountain View, CA).

### Intervention

AFF40 is an annual 40-day alcohol abstinence challenge that takes place in six cities in Louisiana (New Orleans, Covington, Lafayette, Monroe, Shreveport, and Baton Rouge) that begins after the Mardi Gras holiday. The voluntary challenge includes programming to support abstinence, such as webinars, workouts, and a communication campaign, and allows participants to observe changes in their biometric values after abstinence.

### Analysis

Descriptive and inferential analyses were used to examine changes from pre- to post-program participation. Paired t-tests were used to compare pre- and post-program metrics in GraphPad Prism 9.0.1 (San Diego, CA). For percent change calculations, one sample Wilcoxon tests were used to assess whether the median was non-zero.

## Results

### Lab Values

Between pre- and post-AFF40, AST, GGT, total cholesterol, HDL, and triglycerides decreased significantly. The mean differences are shown in **Figure 1**. In 2019, post-program ALT liver enzymes were not significantly different from baseline but significantly dropped by 5.6 U/L in 2020 and 3.6 U/L in 2021. AST significantly dropped in all three years by 2.2-4.8 U/L, and GGT consistently fell, on average, by 7.5-8.7 U/L. Vitamin B12 increased 54.6 pg/mL and 48.7 pg/mL in 2019 and 2021, respectively, but did not significantly change in the 2020 cohort. Total cholesterol dropped by 11.6-15.8 mg/dL across years, HDL dropped by 4.8-8.3 mg/dL, and triglycerides dropped by 26 to 30.2 mg/dL. LDL was unchanged in 2019 and 2020 but decreased by 3.6 mg/dL in the 2021 cohort. For comparison to other studies, the median percent change (rather than absolute change) for each metric is presented in the **Supplemental Figure**.

**Figure 1.**
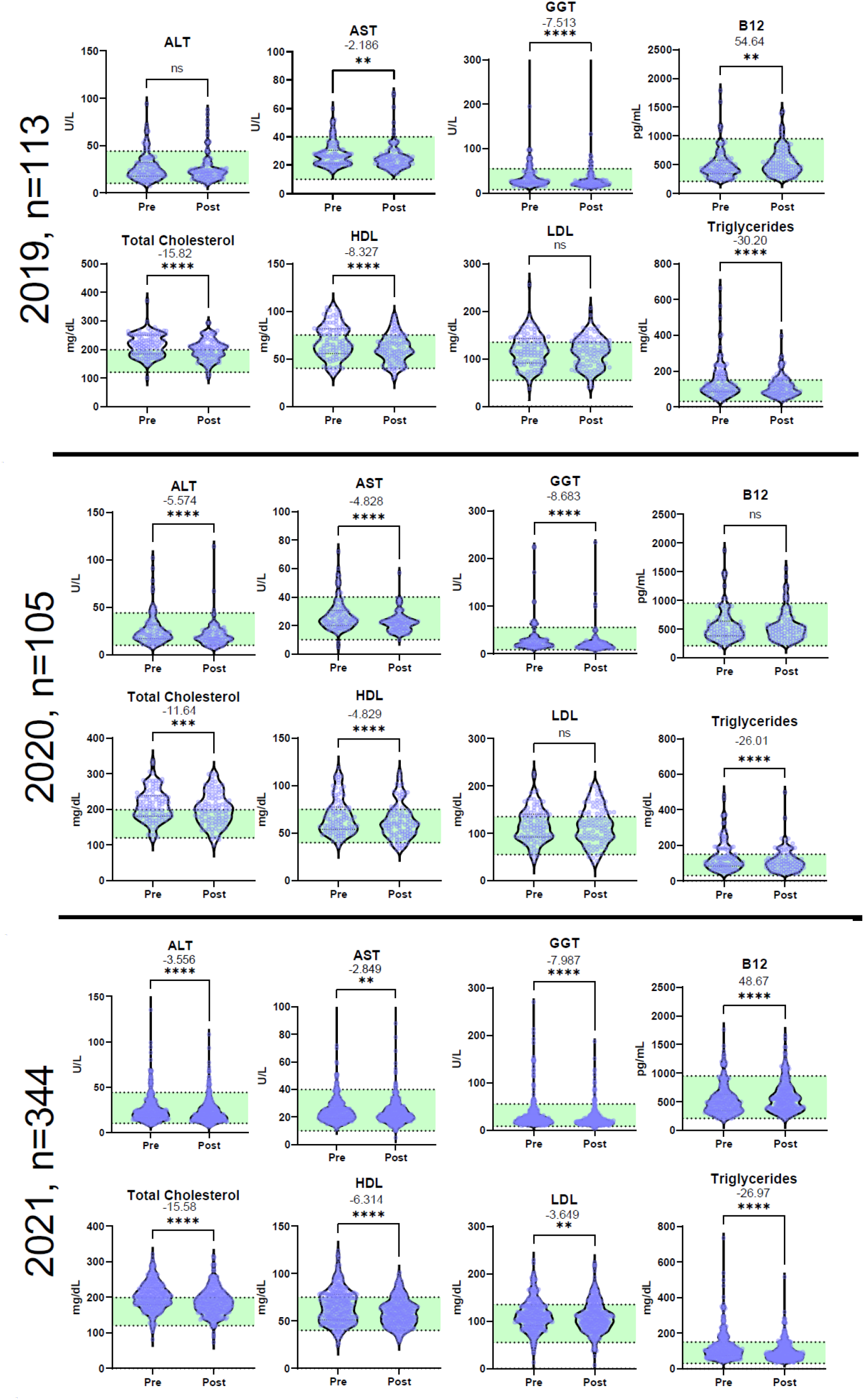
Pre-challenge and post-challenge lab values in 2019 (n=113), 2020 (n=105), 2021 (n=344). Values were separated by 83 days due to the SARS-CoV-2 pandemic in 2020. Paired t-tests were performed and significance (*p<0.05, **p<0.01, ***p<0.001, ****p<0.0001) and difference of means are indicated at the top of each graph. Normal range of values is indicated in green. ALT, Alanine aminotransferase; AST, Aspartate transaminase; GGT, gamma-glutamyl transferase; B12, Vitamin B12; CHOL; total cholesterol; HDL, high-density lipoprotein; LDL, low-density lipoprotein; TRIG, triglycerides.

### Physical metrics

Violin plots showing the distribution, statistical significance, and mean of differences for pre- and post-program are shown in **Figure 2**. Systolic blood pressure decreased by 9 mmHg in 2019 and 7mmHg in 2021 and diastolic blood pressure decreased in 2021 by 4.3 mmHg. Weight, pounds of body fat, and percent body fat all decreased significantly in both years. The mean change in weight (−5.7 lbs, 2019; -3.8 lbs, 2021) is mostly accounted for by the loss of body fat (−4.4 lbs, 2019; -3.3 lbs, 2021). The median percent change by metric is shown in the **Supplemental Figure**.

**Figure 2.**
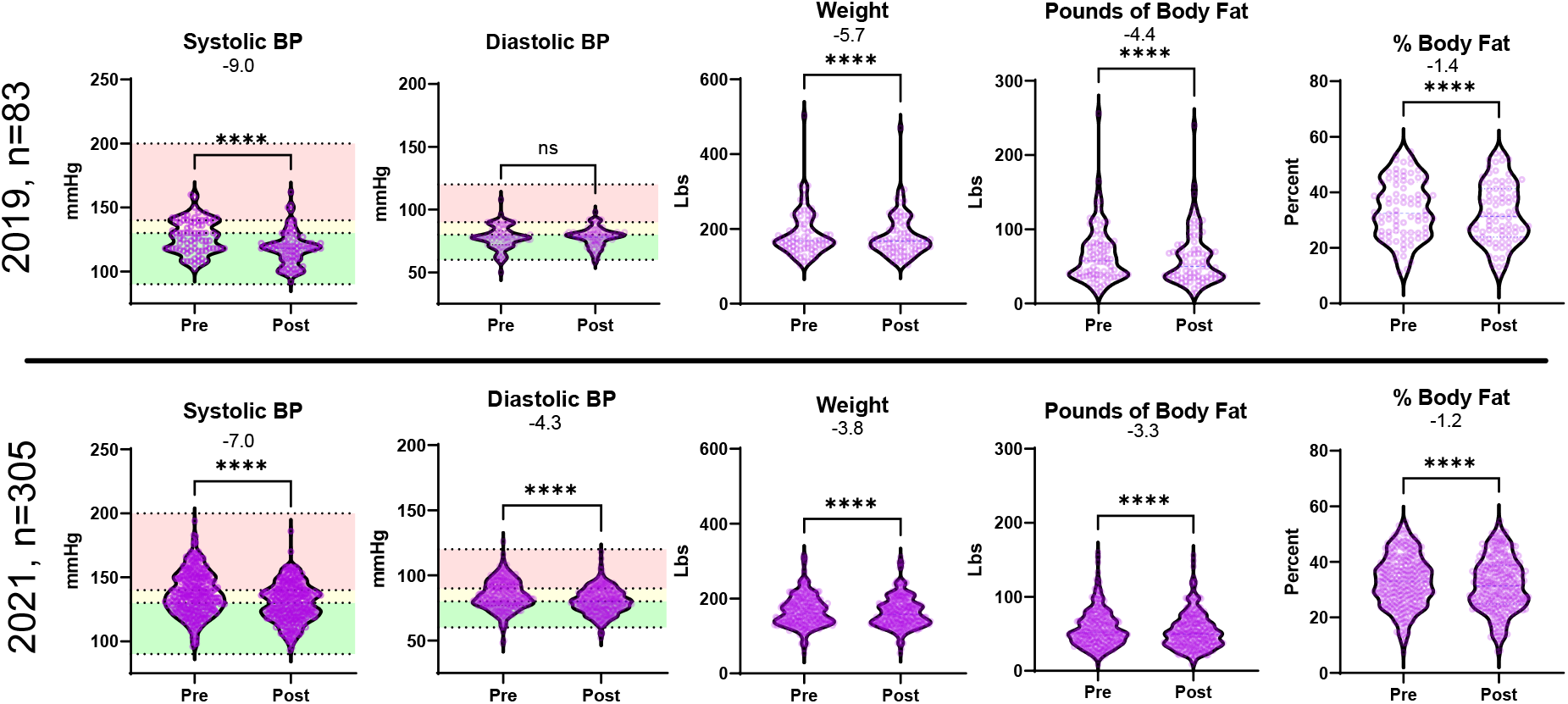
Pre-challenge and post-challenge physical metrics from 2019 (n=83) and 2021 (n=305). Paired t-tests were performed and significance (*p<0.05, **p<0.01, ***p<0.001, ****p<0.0001) and difference of means are indicated directly above each graph. Normal range of values is indicated in green, elevated in yellow, and high in red. mmHg, millimeter of mercury; Lbs, pounds; BP, blood pressure.

## Discussion

This study is one of the few available that measured and observed positive changes in health metrics among participants of a community-based temporary alcohol abstinence program.

Consistent with previous research, AST, ALT, and GGT, liver enzymes released when liver or muscles are damaged, decreased as expected following reduced alcohol consumption.^7^ From pre- to post-challenge, vitamin B12, a micronutrient important in DNA synthesis and neurological function, significantly increased among participants. As alcohol is caustic to the gastric mucosa, the increase in vitamin B12 could indicate better nutrient absorption due to healing of gastric mucosa.^8^

Total cholesterol, HDL, and triglycerides significantly decreased over all three years, and LDL significantly decreased in one cohort. While lipid profiles improved overall, the decrease in HDL is notable as a negative change. Research has shown that light to moderate alcohol intake increases HDL, which may be protective against coronary heart disease; however, heavy drinking, even occasionally, has negative impacts on cholesterol levels.^9^

Group differences were observed in systolic but not diastolic blood pressure in 2019 and significant group differences in both in 2021. A large meta-analysis found that in people who drank more than 2 drinks per day, a reduction in alcohol intake was associated with blood pressure reduction (mean difference systolic: -5.50 mmHg, diastolic: -3.97 mmHg).^10^ Another study of individuals with reduced drinking among those alcohol use disorders noted reduced systolic blood pressure (−4.2 mmHg) but did not record diastolic blood pressure.^7^

In this study, weight and body fat were reduced. Alcohol slows metabolism, can be calorically dense, and can have appetite enhancing effects. Alcohol use may be a risk factor for becoming overweight or obese. ^11^ More research are needed to determine whether the weight and body fat loss were due to reduced alcohol consumption, changes in appetite because of reduced alcohol consumption, or other lifestyle changes (diet and/or physical activity) among participants during the challenge period.

Of note, retention in this study was over 60% in 2019 and over 80% in 2021, which is strikingly better than any intervention across 64 trials reviewed in a meta-analysis of alcohol abstinence interventions.^12^ Even in 2020 during a pandemic when many participants avoided public spaces, 23% of participants returned for follow-up lab work

This study was limited by available data. Other factors, such as medications or changes in diet or physical activity, could have played a role in changing lab values and physical metrics. Importantly, those who completed this intervention were mostly white and female, limiting generalizability.

Individuals engaged in a voluntary alcohol abstinence program utilizing biometric measures during a 40-day challenge effectively improved liver enzymes, cholesterol, body fat, and other factors. Programs like this may hold promise in improving health, but programming should be adjusted to ensure accessibility to non-white, non-female participants. Other alcohol-reduction interventions should consider biometric tracking to improve program adherence and measure effectiveness.

## Data Availability

All data produced in the present study are available upon reasonable request to the corresponding author

## Acknowledgements

The authors thank Alexis Weilbaecher, LDN, RDN and Lauren Hulin, LDN, RDN for placing lab orders, Margaret Haferman for primary data extraction from the electronic medical record, and Lindsey Himmaugh for patient scheduling and coordination. The authors would especially like to thank supervising medical staff for the Alcohol Free for 40 programs, Dr. Dean Hickman, Dr. Michael O’Neal, and Dr. James Patterson.

---

**Supplemental Figure.**
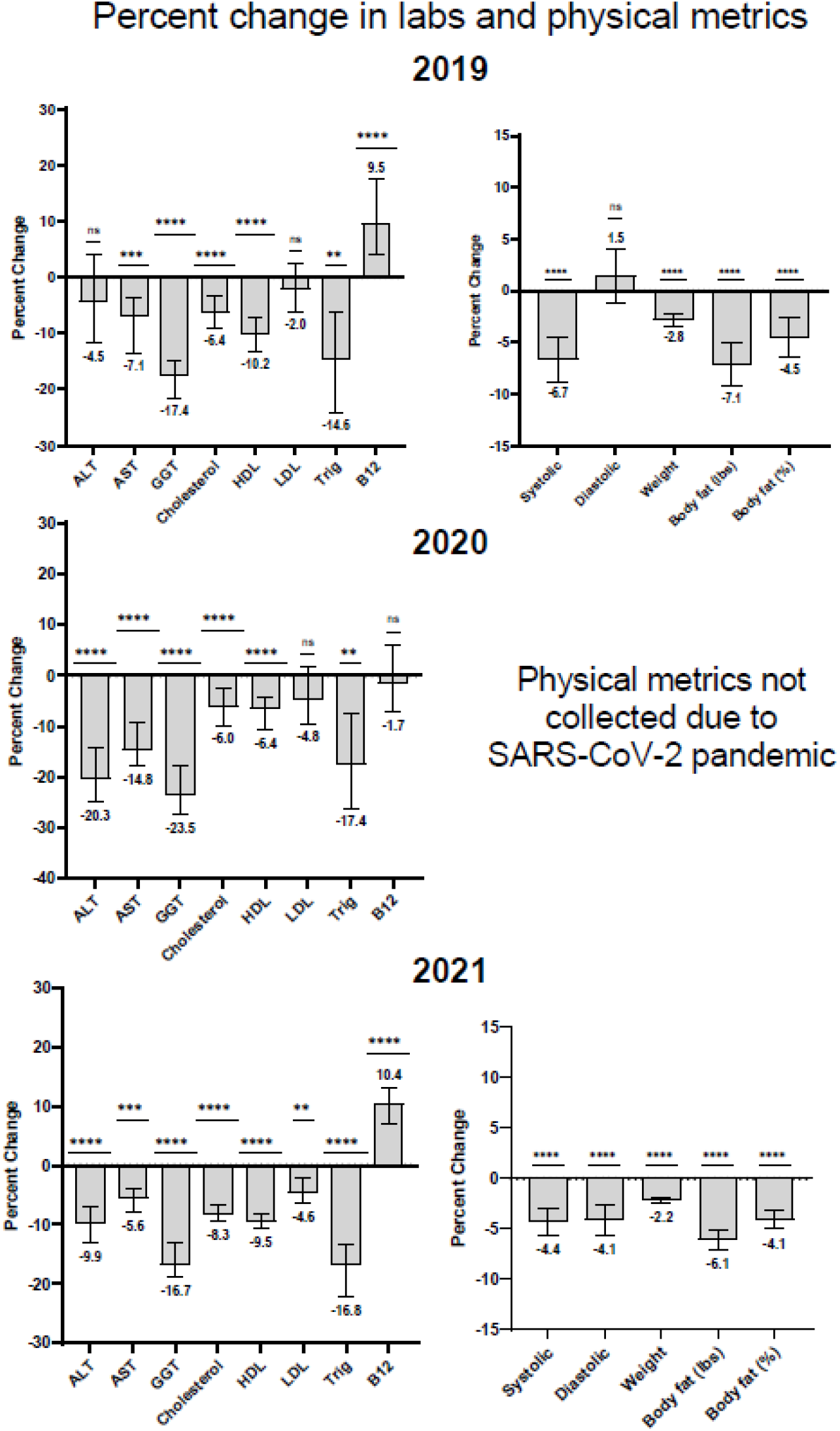
Percent change ([post-pre]/pre) in lab values and physical metrics were calculated for each participant and the median and 95% confidence intervals were graphed. One sample Wilcoxon tests were used to assess the median percent change as non-zero (*p<0.05, **p<0.01, ***p<0.001, ****p<0.0001).

## References

1. Centers for Disease Control and Prevention, National Center for Chronic Disease Prevention and Health Promotion, Division of Population Health. BRFSS Prevalence & Trends Data [online]. 2015. [accessed Feb 11, 2022]. URL: https://www.cdc.gov/brfss/brfssprevalence/.

2. Esser MB, Hedden SL, Kanny D, Brewer RD, Gfroerer JC, Naimi TS. Prevalence of alcohol dependence among US adult drinkers, 2009-2011. Prev Chronic Dis. Nov 20 2014;11:E206. doi:10.5888/pcd11.140329

3. Rehm J, Baliunas D, Borges GL, et al. The relation between different dimensions of alcohol consumption and burden of disease: an overview. Addiction. May 2010;105(5):817–43. doi:10.1111/j.1360-0443.2010.02899.x

4. Global status report on alcohol and health - 2018 (World Health Organization) (2018).

5. Mehta G, Macdonald S, Cronberg A, et al. Short-term abstinence from alcohol and changes in cardiovascular risk factors, liver function tests and cancer-related growth factors: a prospective observational study. BMJ Open. May 5 2018;8(5):e020673. doi:10.1136/bmjopen-2017-020673

6. Munsterman ID, Groefsema MM, Weijers G, et al. Biochemical Effects on the Liver of 1 Month of Alcohol Abstinence in Moderate Alcohol Consumers. Alcohol Alcohol. Jul 1 2018;53(4):435–438. doi:10.1093/alcalc/agy031

7. Witkiewitz K, Kranzler HR, Hallgren KA, et al. Drinking Risk Level Reductions Associated with Improvements in Physical Health and Quality of Life Among Individuals with Alcohol Use Disorder. Alcohol Clin Exp Res. Dec 2018;42(12):2453–2465. doi:10.1111/acer.13897

8. Bode C, Bode JC. Alcohol’s role in gastrointestinal tract disorders. Alcohol Health Res World. 1997;21(1):76–83.

9. Handa K, Sasaki J, Saku K, Kono S, Arakawa K. Alcohol consumption, serum lipids and severity of angiographically determined coronary artery disease. Am J Cardiol. Feb 1 1990;65(5):287–9. doi:10.1016/0002-9149(90)90289-d

10. Roerecke M, Kaczorowski J, Tobe SW, Gmel G, Hasan OSM, Rehm J. The effect of a reduction in alcohol consumption on blood pressure: a systematic review and meta-analysis. Lancet Public Health. Feb 2017;2(2):e108–e120. doi:10.1016/S2468-2667(17)30003-8

11. Traversy G, Chaput JP. Alcohol Consumption and Obesity: An Update. Curr Obes Rep. Mar 2015;4(1):122–30. doi:10.1007/s13679-014-0129-4

12. Cheng HY, McGuinness LA, Elbers RG, et al. Treatment interventions to maintain abstinence from alcohol in primary care: systematic review and network meta-analysis. BMJ. Nov 25 2020;371:m3934. doi:10.1136/bmj.m3934

